# Past SARS-CoV-2 infection elicits a strong immune response after a single vaccine dose

**DOI:** 10.1101/2021.03.14.21253039

**Authors:** Rossana Elena Chahla, Rodrigo Hernán Tomas-Grau, Silvia Inés Cazorla, Diego Ploper, Esteban Vera Pingitore, Mónica Aguilar López, Patricia Aznar, Malena Alcorta, Eva María del Mar Vélez, Agustín Stagnetto, César Luís Ávila, Carolina Maldonado-Galdeano, Sergio Benjamín Socias, Dar Heinze, Silvia Adriana Navarro, Conrado Llapur, Dardo Costas, Isolina Flores, Gabriela Apfelbaum, Raúl Mostoslavsky, Gustavo Mostoslavsky, Gabriela Perdigón, Rosana Nieves Chehín

## Abstract

We hypothesized that in individuals with previous SARS-CoV-2 infection, the first vaccine dose would work as a booster, eliciting a faster and more intense immune response. We herein describe antibody responses to the first and second doses of Gam-COVID-Vac (SPUTNIK V) vaccine in health personnel of Tucumán, Argentina, with previous COVID-19 and compared it with uninfected personnel. Individuals with anti-SARS-CoV-2 titers at baseline showed significantly higher responses to the first dose than people with no prior history of disease (p <0.0001), with titers higher to those registered after the second dose in the control group, representing a clear secondary antibody response. This suggests that a single dose of SPUTNIK V for people with previous SARS-CoV-2 infection could contribute to a better use of available doses.

**One-Sentence Summary:** First vaccine dose in subjects with prior COVID19 elicits a higher antibody response than two doses in uninfected individuals

## Main Text

The need to control the transmissibility and mortality associated with SARS-CoV-2 has transformed vaccines into a critical resource to mitigate the devastating effects of the pandemic originated in Wuham in December 2019 (*1, 2*). The appearance of new virus variants (*3*), whose selective pressure depends on the transmission of the virus, reinforces the need to accelerate the vaccination process in all countries (*4*). However, despite the unprecedented speed of development of different vaccines around the world, their availability remains very limited. This situation is even more critical in underdeveloped or developing countries. In Argentina, the local regulatory authority has so far approved two adenovirus-based vaccines, Gam-COVID-Vac (SPUTNIK V) and Oxford-AstraZeneca, which rely on a prime-booster approach.

The persistence of humoral and cellular immunity against SARS-CoV-2 in convalescent patients has generated new questions regarding the requirement for a two-dose regimen in both adenovirus-based and mRNA vaccines (*5-7*). In this context, this work aimed at studying the post-vaccination humoral immune response to SPUTNIK V in a population of individuals with previous SARS-CoV-2 seroconversion and comparing it with a control group with no previous history of disease.

The immune response to the SPUTNIK V vaccine (rAd26 and rAd5) was analyzed in volunteer healthcare personnel (HCP) who were vaccinated between December 2020 and February 2021 in Tucumán-Argentina. A highly specific and sensitive anti-RBD IgG ELISA test (MEDRXIV/2021/252711, see Methods) was used to study the general response of the HCP population to immunization with SPUTNIK V, following the scheme provided by the Argentinean Ministry of Health. This test detects antibodies raised against the receptor binding domain (RBD) of the Spike (S) protein of SARS-CoV-2, and its correlation with virus neutralization has been well documented (*8, 9*).

Before vaccination, basal IgG anti-RBD titers were measured in 602 HCP volunteers. Antibody titers, calculated as the dilution in which the optical density (OD) obtained was equal to the lowest detectable value that indicates a positive result (see Methods), were detected in 45.2% of volunteers (Fig. 1A, solid and spotted purple), despite the fact that only 21% had confirmed previous natural infection as informed by the public health system (by RT-PCR or rapid antigen test). We consider this population with anti-RBD antibodies at baseline the seropositive group for the purpose of this study. After vaccination with SPUTNIK V, anti-RBD antibody titers were measured at i) 0 days post vaccination (dpv), ii) 14 dpv and iii) 28 dpv (7 days after the second dose), in 252, 520 and 374 samples respectively. At the beginning of the vaccination scheme (0 dpv), the mean IgG anti-RBD titer was 210. After the first dose of SPUTNIK V (14 dpv), the mean titer reached 753, while at 28 dpv, the mean titer rose to 1771, with 97.05% of individuals showing seropositivity (Fig. 1B). A small but statistically significant difference was observed between antibody titers elicited 28 dpv between male and female HCP (Fig. 1C). When subjects were grouped according to age (between 21-60 years of age), no statistical difference was found between mean antibody titers elicited by each age group (Fig. 1D).

**Figure 1.**
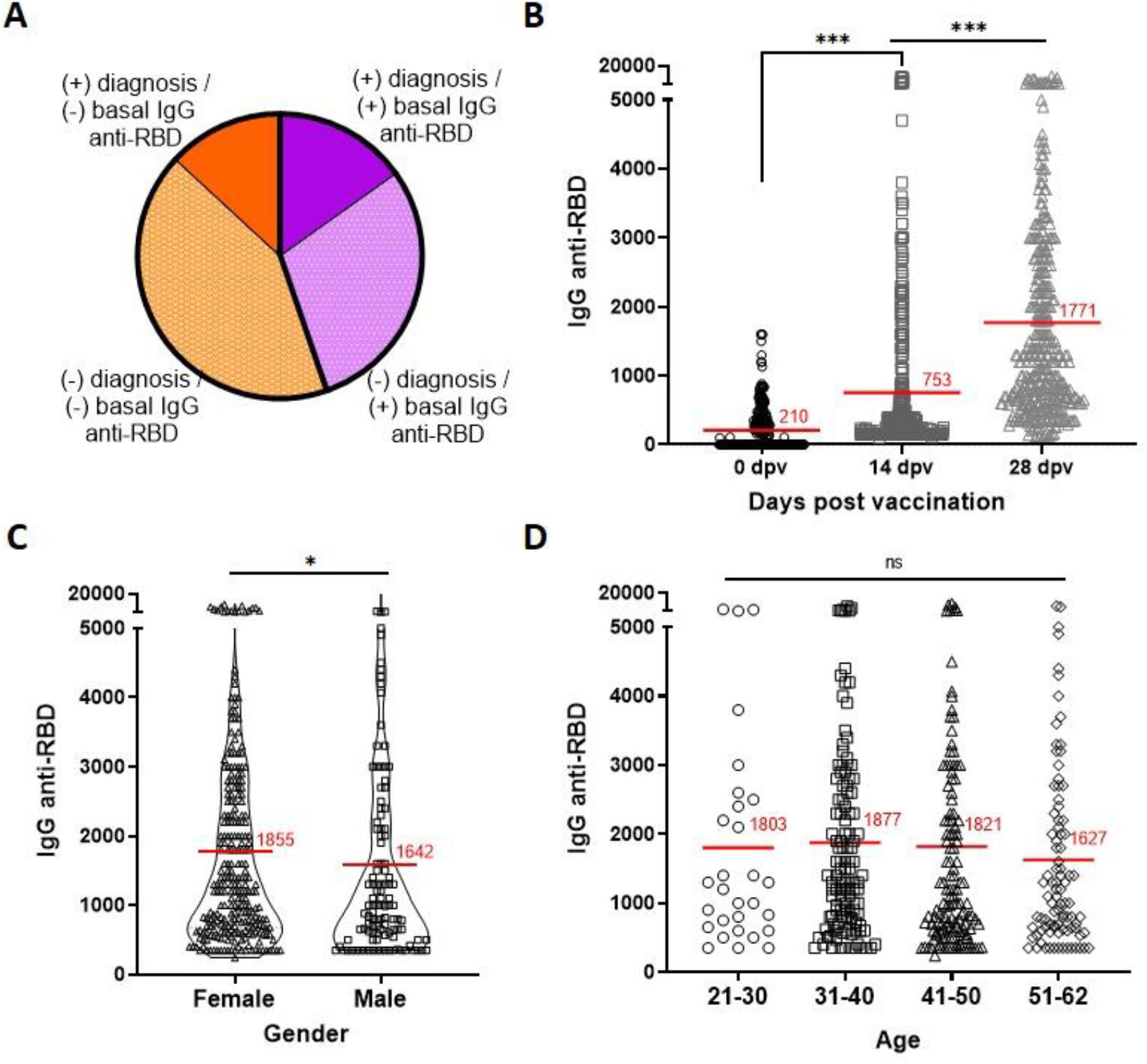
Humoral immune response elicited in 602 individuals after receiving the SPUTNIK V vaccine. (**A**) Distribution of vaccinated population according to the presence of basal IgG anti-RBD and the history of previous documented COVID-19 diagnosis. (**B**) Anti-RBD specific titers, as measured by ELISA, at 0 days post-vaccination (dpv) (black circles), 14 dpv (dark grey squares) and 28 dpv (light grey triangles). *** p<0.0001. Statistical analysis was performed with Dunn’s multiple comparison test. (**C**) Anti-RBD specific titers in female (black triangles) or male (black squares) HCP. * p<0.05. Statistical analysis was performed with Mann-Withney test. (**D**) Anti-RBD specific titers in four defined age groups: 21-30 (circles), 31-40 (squares), 41-50 (triangles), 51-60 (diamonds) at 28 dpv. Mean antibody titer for each group is indicated as a red line. ns=not-significant. Statistical analyses were performed with Dunn’s multiple comparison test.

In order to perform a longitudinal case control study, we followed a group of 214 individuals and analyzed antibody titers measured at 0 dpv and 14 dpv (Fig. 2A). Of these, 107 completed the vaccination scheme with the second dose (Fig. 2B). The first dose of SPUTNIK V (14 dpv), significantly increased antibody titers both in the group with pre-existing SARS-CoV-2 immunity (basal IgG anti-RBD +) (p<0.0001), as well as in those without detectable titers at baseline (p<0.0001) (Fig. 2A). However, the group that was seropositive at baseline achieved a much higher mean antibody titer at 14 dpv (5.4 fold higher) than the control group (p<0.0001). This highly significant difference suggests that a single dose vaccination in previously exposed individuals elicits a secondary-like immune response, acting essentially as an immune boost. Even in individuals who underwent asymptomatic SARS-CoV-2 infection with basal anti-RBD IgG titers, their antibody response was substantially higher at 14 dpv than those with no basal antibodies (5.5 fold higher, p<0.0001) (Fig. S1). This asymptomatic group even registered a small but consistently higher mean antibody titer before being vaccinated (0 dpv) compared to the control group after receiving the first dose (14 dpv) (p<0.0001) (Fig. S1).

**Figure 2.**
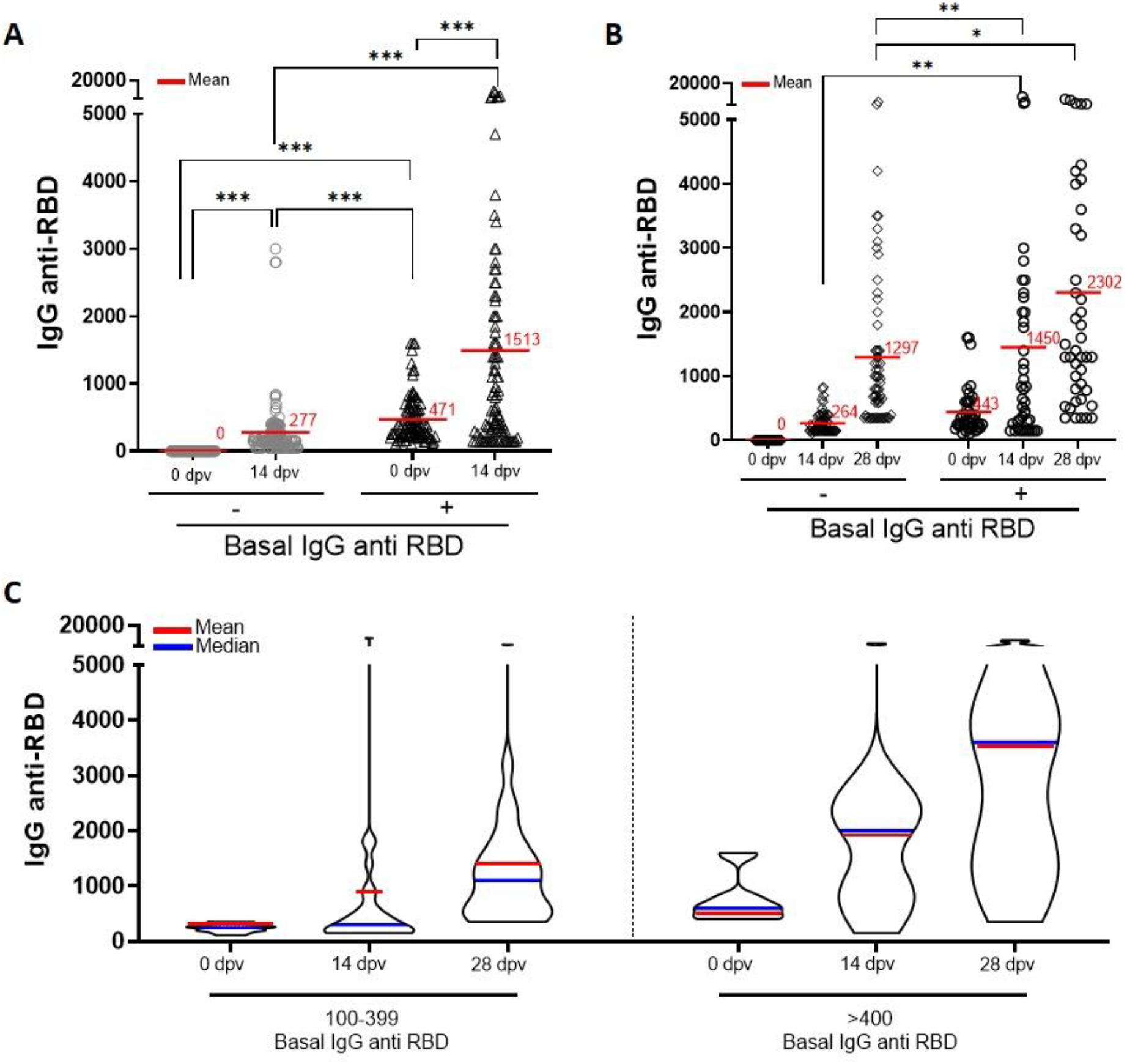
Case and control study of IgG anti-RBD titers upon SPUTNIK V vaccination in individuals with prior SARS-CoV-2 seropositivity compared to uninfected personnel. (**A**) Longitudinal anti-RBD IgG specific titers, as measured by ELISA, at 0 and 14 dpv, in 214 case controlled individuals. (**B**) IgG anti-RBD titers in a subgroup of 107 HCP who completed the full vaccination scheme with the second dose (28 dpv). (**C**) Distribution of previously seroconverted individuals segregated according to basal IgG anti-RBD titers (100-399 and >400) achieved at 0, 14 and 28 dpv in shown as a violin plot. In each panel the mean and the median antibody titer for each group are indicated as a red line or a blue line, respectively. *** p<0.0001 ** p<0.005, *p<0.05. Statistics analysis was performed with Kolmogorov-Smirnov test.

In the subgroup that completed the full vaccination scheme (28 dpv), the second dose of SPUTNIK V elicited higher antibody titers in the group with pre-existing SARS-CoV-2 immunity (2302), compared to the control group (1297) (p <0.05) (Fig. 2B). Notably, a single dose elicited higher antibody titers (14 dpv, 1450) in the seropositive group than two doses of the vaccine in the seronegative group (28 dpv, 1297) (p<0.005) (Fig 2B). Although this difference was statistically significant, the overall distribution of titers within the seropositive subgroup at 14 dpv appeared asymmetric, with the median significantly below the mean, indicative of a highly heterogeneous response to the vaccine (Fig. 2B). Indeed, if this group is further separated based on titers below or above 400, then the response to the first dose becomes strongly homogeneous in >400 subgroup (Fig 2C), strongly suggesting that the anti-RBD IgG titers at baseline have a direct consequence in the overall response to the vaccine. Significant differences in anti-RBD titers were detected after each vaccine dose in both groups, underscoring the general effectiveness of the SPUTNIK V (*10*).

With the aim of determining if there is an optimal window post-SARS-CoV-2 diagnosis in which the SPUTNIK V vaccine stimulates higher antibody titers in seroconverted individuals, we analyzed the effect of days past between diagnosis (in individuals with confirmed positive RT-PCR) and vaccination (days post diagnosis, dpd) on anti-RBD IgG titers. Notably, there was no significant difference associated between dpd (at least up to 120 days) and antibody responses to the vaccine (Fig. 3A-C).

**Fig. 3.**
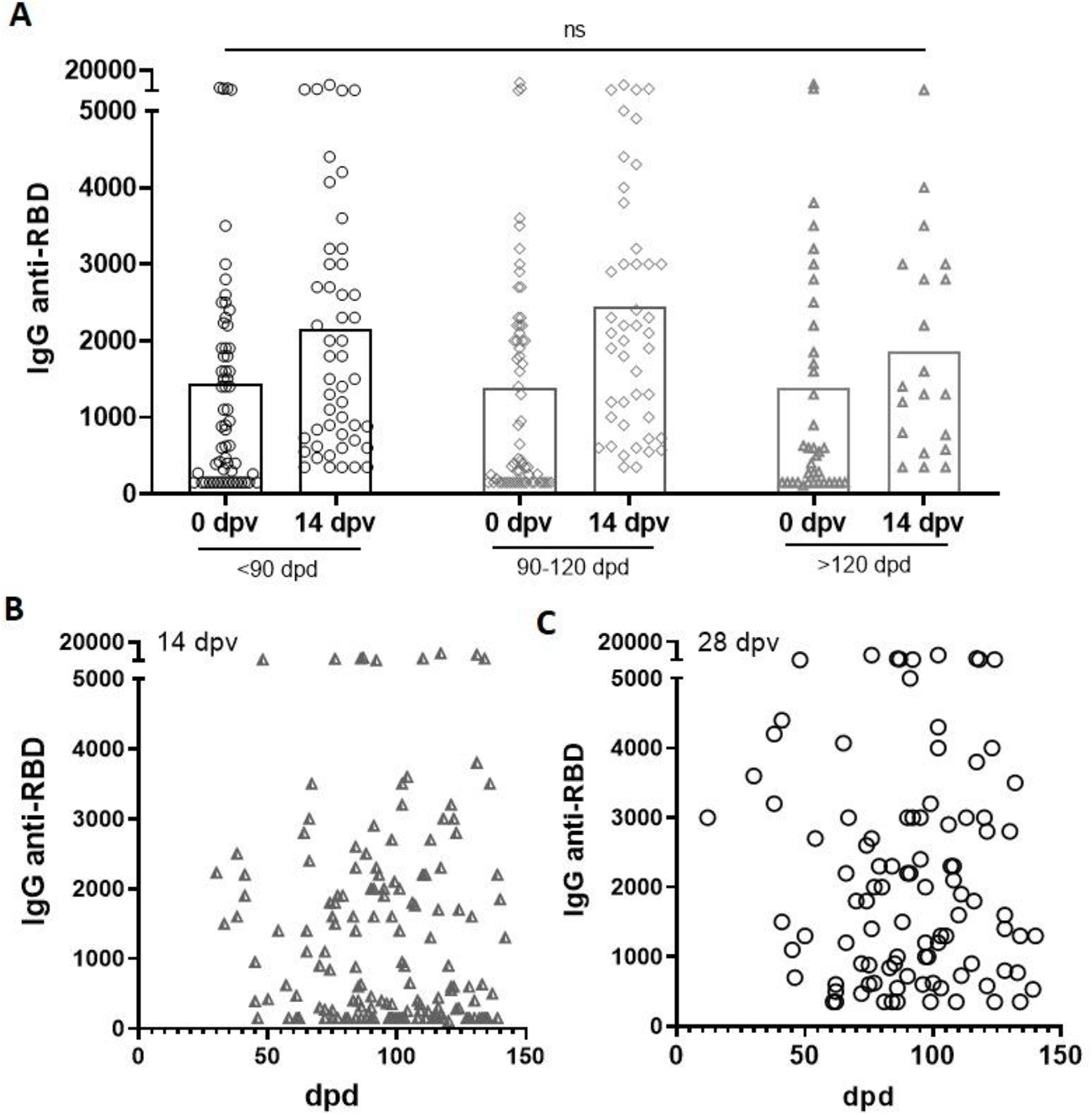
Effect of time elapsed between diagnosis of SARS-CoV-2 and vaccination with SPUTNIK V on anti-RBD antibody titers. (**A**) IgG anti-RBD titers triggered by the first (14 dpv) and second (28 dpv) vaccine doses when administered less than 90, between 90-120, and more than 120 days post diagnosis (dpd) of SARS-CoV-2 infection (by RT-PCR or antigen test) ns=not-significant. Statistical analyses were done with Dunn’s multiple comparison test (**B-C**) Correlation between the dpd and IgG anti-RBD titers generated by the first (B) (r=0.01245, p=0.8770) or second dose (C) of the SPUTNIK V vaccine (r=-0.03879, p=0.7045). The correlation was analyzed using Pearson Correlation Coefficient.

In a context of limited access to SARS-CoV-2 vaccines, our findings show that in individuals previously exposed to the virus, the first dose of SPUTNIK V elicits a strong humoral immune response comparable or even higher than that reached after two doses in individuals without previous exposure to the virus. Our results suggest that the first dose seems to act as a booster for individuals with detectable anti-SARS-CoV-2 antibody titers (regardless if they were asymptomatic or not), and that screening for SARS-CoV-2 seropositive individuals with titers above 400 among the general population could serve to identify those who might require only one vaccination dose.

## Data Availability

all data are available on demand

## Acknowledgments

We thank Mr. Claude Burgio (SkyBio LLC), Ing. Luis Rocha, Ing. Marina Gandur, Dr. Christian Jaroszewski, Dr. María de los Angeles Peral de Bruno and Lic. Lorena Naidicz. We thank Aaron Schmidt for providing valuable reagents.

## Funding

This work was funded by Ministerio de Salud Pública de Tucumán (REC, RTG, SIC, DP, EVP, MAL, PA, MA, EMdMV, AS, CLA, CMG, SBS, DH, SAN, CLL, DC, IF, GP, RNC)

Argentinean Research Council-CONICET grants PIP 722 and 806 (RNC, CMG) Argentinean

Research Agency grants MINCYT PICT-2018-3379 and PICT2018-02989 (DP, SBS)

Tucuman National University grants (PIUNT-UNT D644/1 and D624 (CLA, CMG)

Fundación Florencio Fiorini (DP)

## Author contributions

Conceptualization: RECH, RCH, GP, GA, CLL

Methodology: RHTG, SIC, DP, EVP

Investigation: PA, MA, EMdMV, AS, CLA, SBS, RHTG, DH, SN, CMG

Visualization: RCH, GP, GA, GM, RM

Funding acquisition: RECH, RCH, DP, SBS, CA

Project administration: RCH, MAL

Supervision: RCH, SC, EVP, DC, IF

Writing – original draft: DP, RCH, GP, RHTG, SC

Writing – review & editing: GM, RM, GA, RCH, DP, RHTG, SC, EVP

## Competing interests

Authors declare no competing interests.

## Notes

### Competing Interest Statement

The authors have declared no competing interest.

### Funding Statement

This work was funded by
Ministerio de Salud Publica de Tucuman (REC, RTG, SIC, DP, EVP, MAL, PA, MA, EMdMV, AS, CLA, CMG, SBS, DH, SAN, CLL, DC, IF, GP, RNC)
Argentinean Research Council-CONICET grants PIP 722 and 806 (RNC, CMG)
Argentinean Research Agency grants MINCYT PICT-2018-3379 and PICT2018-02989 (DP, SBS)
Tucuman National University grants (PIUNT-UNT D644/1 and D624 (CLA, CMG)
Fundación Florencio Fiorini (DP)

### Author Declarations

The protocol used in the present study was opportunely submitted to the: -Comite de Etica en Investigacion- Direccion de Investigacion en Salud- SiProSa (Sistema Provincial de Salud). Address: Virgen de la Merced 189-1st floor- (San Miguel de Tucuman) -4000-Argentine. The protocol was approved by Research Department and Ethics Committee (SiProSa-Tucuman Health Ministery). Identification: Dictamen CEI 29-2020 Contact: +54-9-0381-4526585-ext:120 See all information at: https://msptucuman.gov.ar/wordpress/wp-content/uploads/2021/03/Expediente-N3929-410-P2020-Seroconversin-post-vacunacin-para-SARS-CoV-2-immune-response-after-a-single-vaccine-dose.pdf https://msptucuman.gov.ar/wordpress/wp-content/uploads/2021/03/03.Serocoversion-DICTAMEN-19-1-2021.pdf https://msptucuman.gov.ar/wordpress/wp-content/uploads/2021/03/FCI-SeroconversionRCSfinal.pdf

