## Supplementary for "Past SARS-CoV-2 infection elicits a strong immune response after a single vaccine dose"

**This PDF file includes:**

Materials and Methods

Fig. S1

### Materials and Methods

Population studied

Six hundred and two healthcare personnel (HCP) from Tucumán corresponding to 12 different vaccination nodes between December 2020 and February 2021 which received Gam-COVID-Vac (Sputnik V) were invited to participate in the study. The enrolled volunteers who attended the vaccination centers were properly informed regarding their choice to participate in the study. The protocols were approved by the regional ethical research boards (Ex. N° 3929-410-P-2020) following the Declaration of Helsinki. Personal data from all volunteers were encrypted. Eligibility criteria were age between 18 to 60 years asymptomatic to COVID-19 at the time of vaccination. Individuals provided a signed informed consent to be included in the database of the present study.

ELISA anti-RBD antibody assay

IgG anti-RBD titers were determined by using the enzyme-linked immunosorbent assay (ELISA) developed and validated in our laboratory (MEDRXIV/2021/252711). Briefly, recombinant RBD from SARS-CoV-2 was obtained from HEK293 cells, which were transduced with a pHAGE2 lentivirus (pHAGE2-RBD-His) in order to generate a stable transgenic cell line. RBD protein domain was purified by affinity chromatography. Purity was controlled on an SDS-PAGE gel. Purified RBD was immobilized in each well (0.1 µg) of a 96-well flat polystyrene bottom plates (High Binding, Half-Area, Greiner 675061). The ELISA for the determination of IgG anti-RBD presented high sensitivity (92.2%) and specificity (100%). Antibody titer value representing the highest accuracy (Sensitivity + Specificity) was calculated using receiver operating characteristic curve (ROC) to establish the cutoff value. To this end, we used samples from 52 individuals that had positive diagnosis both using RT-PCR amplification of SARS-COV-2 and the Chemiluminescent microparticle immunoassay (CMIA-Architect, Abbot) as positive controls and 26 sera collected before December of 2019 as negative controls. Titers were calculated as the dilution in which the optical density ($OD_{450nm}$) obtained was equal to the cutoff.

Blood samples for IgG anti-RBD detection were taken a) the same day before the first dose application (0 dpv), b) 14 ± 1 days after first dose (14 dpv), c) 28 ± 2 days after first dose (28 dpv). After peripheral blood extraction (5 ml), serum was obtained by spontaneous coagulation in appropriate tubes within two hours of collection. The samples were stored at -20 $^{o}$C.

Statistical analysis

Statistical analysis was performed using non-parametric tests in Prism 8.0 software (GraphPad, San Diego, CA). The post-tests applied in each trial are indicated in the figure captions.

**Figure S1.**

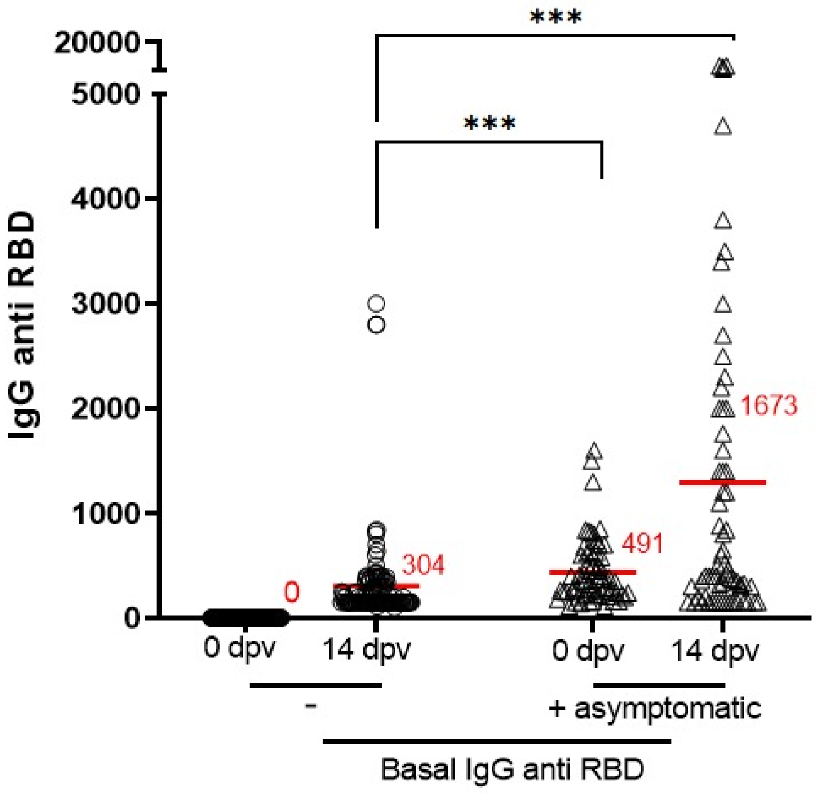


**Fig. S1. Anti-RBD IgG titers triggered by SPUTNIK V first dose in basal seronegative or asymptomatic seropositive individuals.** Anti-RBD IgG antibody titers were determined by ELISA. Samples were taken the day of the vaccine administration (0 dpv) and 14 days after the first dose (14 dpv) of the SPUTNIK V in individuals without prior SARS-CoV-2 infection and in asymptomatic seroconverted individuals. *** $p<0.0001$. Mean antibody titer for each group is indicated as a red line. Statistical analysis was performed with Kolmogorov-Smirnov test.
